# Safety and Preliminary Effectiveness of a Natural, Multi-modal Therapeutic in Postmenopausal Women with Obesity

**DOI:** 10.1101/2025.09.04.25335119

**Authors:** Taylor Thompson, Vineeta Tanwar, William Hoskinson, Kristin Peterson, Melinda Sothern, Christine L. McKibbin, Celestin Missikpode, Pankaj Kapahi, Sanjay Dhar

**Affiliations:** Hoskinson Health and Wellness Clinic, Gillette, WY, United States; Buck Institute for Research on Aging, Novato, CA, United States; Louisiana State University Health Sciences Center-New Orleans, New Orleans, LA, United States; University of Wyoming Center on Aging, Laramie, WY, United States; University of Illinois-Chicago, Chicago, IL, United States

## Abstract

**Introduction:** Postmenopausal women exhibit increased central obesity due to aging-related alterations in hormones leading to adverse metabolic and biological consequences. The present study aimed to evaluate the safety of a natural, multi-modal therapeutic and observe alterations in metabolic and aging-related outcomes in humans for the first time.

**Methods:** A non-comparer pilot study was conducted to translate preclinical findings of a glycation-lowering supplement (Alpha-lipoic acid, nicotinamide, thiamine, piperine, pyridoxamine) to postmenopausal women with obesity [n=85; >55 years; BMI=35.0±4.35/30.3–42.8 (Figure 1)]. Qualified participants (n=13) consumed two capsules daily for 6 months. Complete Blood Count [e.g. (Red Cell Distribution Width (RDW%)], Mean Corpuscular Volume (MCH), etc.], depression (Center for Epidemiologic Studies Depression Scale), insulin resistance [Homeostatic Model (HOMA-IR)], fat oxidation [respiratory quotient (RQ)], weight/height, waist circumference (WC), bone mineral density (BMD) by Dual-Energy X-ray Absorptiometry, low (LDL) and high density (HDL) lipoproteins, phenotypic age (PA), follicle stimulating hormone (FSH) caloric intake [(CI)-NIH/NIS/ASA 24-2020 Dietary Assessment], and immediate recall [(IM) BrainCheck] were assessed at baseline and 6-month-follow-up.

**Results:** No serious adverse events were reported. Six participants reporting mild/moderate adverse events were lost to follow-up. Mixed-effect models (intent-to-treat analysis) compared outcomes prior to (n=13) and following (n=7) the 6-month intervention. RDW% (p=0.009), MCV (p=0004), RQ (p=0.02), WC (p=0.02), HDL (p=0.044), phenotypic age (p=0.037), FSH (0.002), and CI (p=0.01) significantly decreased. Depression (p=0.002), height (p=0.003), BMD (p=0.02) and IM (p=0.04) significantly increased; HOMA-IR and LDL were unchanged.

**Conclusions:** Preliminary results indicate that a natural, multi-modal therapeutic is safe in postmenopausal women with obesity. Several metabolic and aging-related outcomes improved following the intervention. However, decreased HDL, RDW%, MCV and increased depression warrant further investigation in future randomized-controlled trials. Clinicaltrials.gov #NCT06242535

## 1 Introduction

Metabolic and biological aging pathologies are well-established consequences of excess adiposity in older adults affecting more than 764 million worldwide.^1^ Central obesity, in particular, is both a clinical manifestation and driver of metabolic, physical and cognitive aging-related debility, each with similar incidence rates.^2^ Postmenopausal women are especially at increased risk of developing central obesity due to biological aging alterations in endogenous ovarian hormones.^3^ The resulting aging-related metabolic and biological consequences are manifested as impaired fat oxidation [Respiratory Quotient (RQ)],^4^ increased insulin resistance (IR) and glycosylated hemoglobin (HbA1C), reduced lean muscle and bone mass,^5^ physical frailty^6^ and cognitive decline. Unfortunately, current therapeutic methods to address these complex aging pathologies lack specificity and propose universal solutions to a multi-factorial disorder requiring a precision medicine approach.^7^ Pharmacologic interventions, such as Glucagon-like peptide-1 (GLP-1) receptor agonists, are plagued by side effects^8,9^ and high costs to patients.^10^ Currently, there is a lack of rigorous scientific evidence to support conventional dietary therapies in older women specifically.^6^ Non-traditional approaches, such as nutrition therapeutics, are emerging.^11,12^ However, recent studies focus on a solitary ingredient and its effect on a specific biological pathway.^13^ A recent Task Force of experts in geroscience recommend that the current health paradigm alter its focus from the treatment of a single disease to the promotion of therapies that address multiple mechanisms of aging pathology.^7^

Advanced glycation end products (AGEs) are generally formed during metabolism. These AGEs accumulate with age and lead to glycation stress that drives IR, inflammation and oxidative stress, key mechanisms of metabolic dysfunction. Postmenopausal women are particularly susceptible to metabolic dysfunction,^3^ which may partially be driven by elevated AGEs.^14^ A glycation-lowering natural, oral supplement, commercially available as GLYLO, was developed that operates synergistically to detoxify methylglyoxal (MGO), a reactive precursor to AGEs. A by-product of glycolysis, MGO is known to promote obesity,^15^ overeating behavior, neuronal damage and shortened lifespan.^16^ This formulation is composed of five compounds, 1) Alpha lipoic acid (ALA), 2) nicotinamide, 3) thiamine, 4) piperine and 5) pyridoxamine (Table 1). Each of these target distinct metabolic and brain health pathways, and extend healthspan via a mechanism that supports neuronal resilience to methylglyoxal (MGO) in culture.^17–23^ The compounds are capable of producing synergistic effects because each function through a different metabolic and biological aging pathway. Our preclinical findings indicate that supplementation with this natural, multi-modal therapeutic lowered serum MGO levels in diabetic mice, reduced IR, calorie intake (CI), body weight, and central obesity, improved glucose tolerance, and increased physical activity, neuro-muscular function, and lifespan in older, wild-type mice [equivalent to 69-year-old humans (unpublished data available as a preprint].^24,25^ Further research revealed that glycation stress contributed to aortic stiffening, a condition mitigated by glycation stress-lowering compounds in aged mice (unpublished data available as a preprint).^24^ Additionally, in female ovariectomized mice, the treatment reduced the serum FSH levels (unpublished data), a biomarker indicative of improved ovarian function in postmenopausal women.

**Table 1:** Pharmacokinetic Safety Profile of the Natural, Multi-modal Therapeutic Compounds: mg (milligrams)

|  |  |  |  |  |  |  |
| --- | --- | --- | --- | --- | --- | --- |
| Natural Multi-modal Therapeutic Compounds | Metabolism | Route of Elimination | Half Life | Clinically Recommended Dose (mg/day) | Maximum Safety Dose Tested (mg/day) | Study Treatment Dose (mg/day) |
| Pyridoxine (Vitamin B6) | Liver | Urine | 15-20 days | 100-200 | 200 | 50 |
| Thiamine (Vitamin B1) | Liver | Urine | 14-18 days | 50-100 | 500 | 100 |
| Nicotinamide (Vitamin B3) | Liver | Urine | 7-9 days | 100-500 | 900-1500 | 200 |
| Alpha Lipoic Acid | Liver | Urine | 0.5 to 1 hour | 100-300 | 1000-1800 | 150 |
| Piperine | Liver | Urine; Biliary | 1-2 hours | 5-20 | Up to 50 | 10 |

Numerous clinical studies have investigated the effect of the individual ingredients included in this natural, multimodal therapeutic.^13,17,26,27^ Yet, presently there are no clinical studies evaluating the rigorously-tested combination of glycation lowering GRAS (Generally Regarded As Safe) ingredients, which are needed to validate promising preclinical findings. The objective of this pilot study was to translate preclinical findings^15,16,24,25^ into humans by examining the safety and observing the preliminary efficacy of this natural, oral multimodal therapeutic in a cohort of postmenopausal women with obesity ≥ 30 Body Mass Index (BMI). The study also explored multiple aging-related, metabolic and biological pathways related to preclinical findings with novel diagnostics through an integrated, precision medicine approach. Exploratory outcomes were utilized to establish a metabolic and aging-related phenotype specific to female adults. We hypothesized that following six-months of daily supplementation, a natural, multimodal therapeutic^24^ supplementation would demonstrate safety, decrease RQ, IR, HbA1C, waist circumference, fat mass (FM), FSH, CI and phenotypic age,^2^^8^ while increasing lean mass (LM), bone mineral density (BMD), physical (PF) and cognitive function in this cohort of postmenopausal women with obesity.

## 2 Materials and Methods

### 2.1 Patient and Public Involvement

Based on patients’ comments during the phone screening interviews, the original study protocol was amended to reduce study participant burden. The total number and frequency of study assessments was reduced in the study design.

### 2.2 Study Design

A single-center, non-comparer pilot study was conducted to determine the safety, tolerability and preliminary efficacy of six months of an orally delivered natural, multimodal therapeutic in decreasing metabolic disease risk (RQ, IR, HbA1C) and improving BMI and body composition, e.g., LM, FM, BMD, waist circumference, (primary outcome variables) in postmenopausal women with obesity (Figure 1). Secondary metabolic and biological aging-related outcomes of interest were also explored including FSH, depression, CI, phenotypic age, PF, cognitive function and others. Retinal imaging technology was used to apply general aging clock procedures (eyeAge) to explore study participants’ age gap. Study procedures were completed with approval and oversight by the Biomedical Research Alliance of New York (BRANY) Institutional Review Board (IRB). To reduce study participant burden, an amendment was submitted and approved by the IRB prior to the implementation of the baseline assessments as follows: 1) The 3-month assessment clinic visit was removed from the protocol, 2) Blood tests to measure growth hormone, insulin-like growth factor-1, estrone, testosterone, and dehydroepiandrosterone and all inflammatory biomarkers with the exception of C-reactive protein were removed from the protocol. 3) Blood tests to observe adherence by examining the oral therapeutic’s ingredients were removed from the protocol and replaced with pill counts and interviews, 4) Pulmonary function testing was removed from the protocol, and 5) The 4-month ECG assessment was removed from the study protocol. Following IRB approval of the amendment, study assessments were conducted during clinic visits prior to and six months following the intervention. Additional safety assessments were conducted during clinic visits at two and four months during the intervention; the ECG was conducted at the two month visit only (Figure 1).

**Figure 1.**
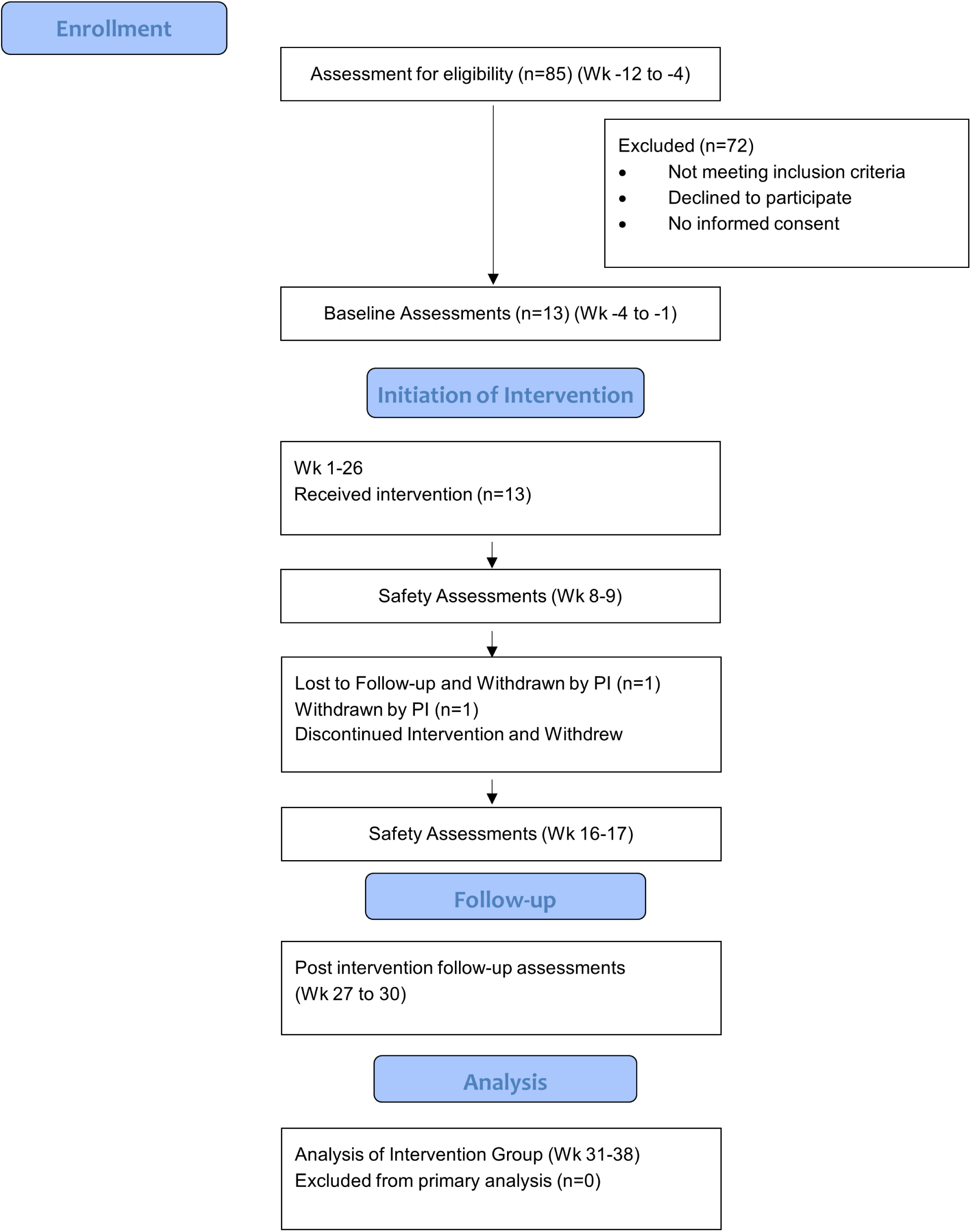
Flow of Participants through Each Stage of the Pilot Clinical Trial (CONSORT Flowchart)

### 2.3 Study Population

Healthy, postmenopausal (> 1 year from last menstrual cycle) adult females > 55 years of age with obesity (BMI ≥ 30) were enrolled at the Hoskinson Health and Wellness Clinic (HHWC), located in Gillette, Wyoming. The HHWC utilizes translational geroscience^29^ and integrated, precision medicine to identify pathological aging biomarkers, epigenetic vulnerabilities, and innovative medical technology to both diagnose and manage aging-associated declines in metabolic, cognitive, and physical health. State-of-the-art assessments identify targets for tailored programming such as nutrition supplementation.

### 2.4 Treatment

A combination of GRAS compounds, the oral supplement is FDA-approved for weight loss and anti-aging (IND 162006) and is commercially available for over-the-counter purchase.^30^ Tishcon corporation manufactured and provided the certificate of analysis of the active supplement pills.^30^ Each participant consumed two capsules per day orally with breakfast in the morning between 7:00–11:00 AM for 6 months. Doses were based on the dose conversion from mice to humans and prior safety data for each of the compounds in humans (Table 1).

### 2.5 Recruitment, Screening, Selection and Withdrawal of Study Participants

Recruitment, screening and enrollment were employed including disseminating flyers and social media posts. Screening interviews and baseline assessments were implemented over a 4-week window prior to intervention initiation. A total of 85 postmenopausal women self-reporting obesity participated in a telephone screening interview to determine eligibility. Participants verbally self-reported information to the interviewer to complete a check list of established inclusion and exclusion criteria for the study. Nineteen of these women met the criteria and were enrolled into the study (BMI=35.0±4.35; Range: 30.3–42.8). Trained clinical research staff obtained informed consent from all participants and obtained detailed medical histories to determine eligibility.

### 2.6 Inclusion Criteria

Postmenopausal (> 1 year from last menstrual cycle) adult females (at birth), > 55 years of age with OW/OB (BMI ≥ 30)

### 2.7 Exclusion Criteria

Adult males including those not female at birth were excluded. Adult females with BMI <30, surgical menopause or receiving hormone replacement therapy (HRT) and pregnant or nursing mothers were excluded. Adult women diagnosed with severe asthma (American Thoracic Society (ATS) standards and/or NIH guidelines), an acute lower respiratory or GI infection within 2 weeks, moderate to severe cognitive impairment, gout, an eating disorder, cardio-metabolic disease [e.g., cardiovascular disease (CVD), type 2 diabetes (T2DM), hypertension, h/o stroke etc.], other chronic, immune, pulmonary, neurodegenerative or systemic disease requiring prescription medication were excluded. Adult women receiving monoclonal antibody or biological treatment or currently prescribed or received weight loss medications within the past 6 months, or currently enrolled in a defined weight loss program were also excluded. Those reporting moderate to severe mental or physical disability or unwillingness to travel to clinic site facilities were excluded. Adult women receiving blood thinners and anti-platelet drugs (other than aspirin), chronic steroids (including inhaled), spironolactone, statins, diuretics, chronic pain medications, or any medication affecting glucose, insulin, lipids, or metabolic performance were also ineligible. Lastly, individuals with recent change in chronic disease, change in medication within the past three months, or those receiving supplements with potential to augment or compete with the natural, multi-modal therapeutic were excluded.

The following medications were allowed before and during the trial if the study participant self-reported that they were stable for >6 months with no recent dose changes: antidepressants (e.g., SSRIs, SNRIs, SARIs, NDRIs, TCAs), ADHD medication (psycho-stimulants), antihistamines, protein pump inhibitors, histamine H2-receptor antagonists (H2 Blockers). Allowable supplements permitted before and during the trial included Magnesium, Omega-3, Probiotics, Digestive enzymes, Collagen, SAMe, Curcumin.

### 2.8 Methods

Screening/Baseline Evaluation and Follow-up Assessments: Following the IRB-approved consent procedures, a screening visit obtained medical history, physical exam including electrocardiogram (ECG), blood tests, and direct measures of weight and height to calculate BMI. These assessments were conducted to confirm eligibility and establish baseline values for safety and tolerability. Participants also completed a Medical Symptoms Questionnaire (MSQ). Study participants were scheduled to attend the following assessments and time points: 1) A safety assessment including blood tests and the MSQ at 2-, 4-, and 6-month follow-up visits; 2) an ECG at baseline and 2 months and; 3) A follow-up assessment consisting of a 2-day visit after 6 months of receiving the oral supplement to assess safety and study outcomes and to observe the preliminary effectiveness of the intervention.

Primary and secondary outcomes were obtained over 2 days at baseline and 6-month follow-up visits. Safety and compliance assessments were conducted at 2-, 4- and 6-month visits by trained technicians. Postmenopausal women (n = 85; > 55 years) were assessed for eligibility. Those eligible (N = 19) were screened; however, six were excluded during the screening visit because they did not meet the inclusion criteria for BMI ≥ 30 when height and weight were measured. A total of 13 (Mean ± SD = 35.0 ± 4.35; Range: 30.3 – 42.8; Table 2) met eligibility criteria and were enrolled into the study. Of these, 13 were clinically defined with obesity [BMI ≥ 30 - (U.S. CDC)] and 7 of the participants with significant obesity (BMI ≥ 35). Prior to and following the 6-month intervention, participants were assessed for the following parameters:

**Table 2:** Baseline Descriptive Characteristics of the Study Participants. SD (standard deviation); BMI [Body Mass Index = total body weight (kg)/Height (m)^2^ (BMI)]; SPB (Systolic Blood Pressure); DBP (Diastolic Blood Pressure; BMD (Bone Mineral Density); BMC (Bone Mineral Content); VAT (Visceral Adipose Tissue); HOMA-IR (Homeostatic Model for Insulin Resistance); hbA1C (glycosylated hemoglobin); TG (Triglycerides); TC (total cholesterol); HDL (High Density Lipoprotein); LDL (Low Density Lipoprotein); CRP (C-Reactive Protein); Alb (Albumin); RDW (Red Cell Distribution Width); LYM (Lymphocytes); CREAT (Creatine) MCV (Mean Cell Volume); ALP (Alkaline Phosphatase); WBC (White Blood Cells); RQ (Respiratory Quotient); Submax [Submaximal (80% heart rate max)] VO_2_ (Oxygen Uptake); MMSE (Mini Mental State Examination of Cognitive Function); BCCM (BrainCheck Cognitive Measurement); BCSS (BrainCheck subscale); CES-D (Center for Epidemiologic Studies Depression Scale); eyeAGE (Retina Scan: General Aging Clock Retina Imaging)

| Baseline Descriptive Characteristics of the Study Participants |  |  |  |
| --- | --- | --- | --- |
| Characteristics | All Participants<br>(n = 13)<br>Mean (SD) | Completers<br>(n = 7)<br>Mean (SD) | Dropouts<br>(n = 6)<br>Mean (SD) |
| Age (years) | 62.05 (3.95) | 61.14 (3.35) | 63.12 (4.64) |
| Weight (kg) | 92.2 (13.68) | 89.2 (9.97) | 95.6 (17.40) |
| Height (cm) | 162.1 (3.97) | 162.4 (3.57) | 161.8 (4.71) |
| BMI | 35.0 (4.35) | 33.8 (3.81) | 36.4 (4.90) |
| SBP | 140.9 (15.24) | 135.9 (14.35) | 146.8 (15.32) |
| DBP | 87.3 (9.41) | 84.9 (8.63) | 90.2 (10.25) |
| Waist Circumference (cm) | 112.5 (12.64) | 111.3 (6.66) | 114.0 (18.03) |
| Physical Function (Total) | 11.5 (0.52) | 11.7 (0.49) | 11.3 (0.52) |
| Fat (%) | 43.9 (2.89) | 42.3 (2.02) | 45.7 (2.74) |
| Fat Mass (kg) | 40.6 (8.29) | 37.7 (5.10) | 44.0 (10.40) |
| Lean Mass (kg) | 51.4 (5.76) | 51.3 (4.63) | 51.5 (7.34) |
| BMD (g/cm<sup>2</sup>) | 1.0 (0.13) | 1.0 (0.11) | 1.1 (0.15) |
| BMC (g) | 2061.5 (364.46) | 2090.8 (290.67) | 2027.4 (463.47) |
| Vat Area (cm<sup>2</sup>) | 195.9 (54.46) | 191.7 (39.89) | 200.8 (71.80) |
| VAT Volume (cm<sup>3</sup>) | 1022.8 (283.76) | 1000.7 (208.15) | 1048.5 (373.87) |
| Vat Mass (g) | 946.2 (262.46) | 925.7 (192.70) | 970.0 (345.68) |
| HOMA-IR | 2.4 (1.12) | 2.3 (1.25) | 2.5 (1.09) |
| hbA1C (%) | 5.5 (0.48) | 5.7 (0.24) | 5.4 (0.61) |
| FSH (mIU/mL) | 65.3 (22.84) | 59.1 (21.08) | 71.5 (24.76) |
| TG (mg/dL) | 192.7 (107.93) | 249.8 (128.16) | 135.5 (36.84) |
| TC (mg/dL) | 248.3 (28.59) | 265 .0 (30.62) | 231.5 (13.66) |
| HDL (mg/dL) | 63.3 (14.82) | 55.2 (13.26) | 71.3 (12.27) |
| LDL (mg/dL) | 146.3 (21.53) | 159.7 (22.84) | 133.0 (8.46) |
| TSH (uIU/mL) | 2.4 (1.22) | 2.8 (0.66) | 2.2 (1.50) |
| CRP (mg/dL) | 0.7 (0.53) | 0.6 (0.21) | 0.7 (0.76) |
| Alb (g/dL) | 4.5 (0.39) | 4.7 (0.44) | 4.3 (0.27) |
| RDW (%) | 12.9 (0.68) | 13.0 (0.76) | 12.7 (0.62) |
| LYM (%) | 35.8 (7.97) | 35.7 (7.21) | 35.8 (9.36) |
| CREAT (mg/dL) | 0.9 (0.20) | 1 (0.25) | 0.8 (0.06) |
| MCV (fL) | 94.2 (2.34) | 93.8 (1.96) | 94.6 (2.80) |
| ALP (U/L) | 82.5 (12.01) | 85.7 (13.87) | 79.3 (10.05) |
| WBC (K/uL) | 5.8 (1.08) | 6.2 (0.68) | 5.5 (1.36) |
| Phenotypic Age | 57.5 (6.47) | 59.0 (5.48) | 56.0 (7.53) |
| RQ | 0.8 (0.06) | 0.8 (0.06) | 0.8 (0.06) |
| Submax VO<sub>2</sub> (ml/min) | 1408.3 (289.38) | 1372.3 (347.16) | 1450.3 (228.94) |
| Submax VO<sub>2</sub>/kg (ml/min/kg) | 15.2 (2.54) | 15.3 (2.87) | 15.1 (2.36) |
| Lean Adjusted Submax VO<sub>2</sub> | 27.4 (4.63) | 26.6 (5.46) | 28.3 (3.69) |
| RQ during exercise | 0.9 (0.07) | 0.9 (0.06) | 0.9 (0.07) |
| Caloric Intake | 1875.7 (545.63) | 1984.9 (569.88) | 1748.4 (537.27) |
| MMSE Total (score) | 29.5 (0.53) | 29.5 (0.59) | 29.6 (0.49) |
| BCCM Total (score) | 100.8 (8.82) | 100.9 (9.53) | 100.7 (8.82) |
| BCSS Trails A (score) | 107.6 (4.84) | 106.0 (3.37) | 109.5 (5.89) |
| BCSS Trails B (score) | 104.6 (7.76) | 105.5 (6.35) | 103.7 (9.50) |
| BCSS Digit (score) | 103.1 (10.68) | 103.4 (12.03) | 102.7 (9.99) |
| BCSS Stroop (score) | 105.4 (9.74) | 103.6 (9.48) | 107.5 (10.48) |
| BCSS Immediate Recall (score) | 96.6 (14.61) | 98.0 (19.32) | 95.0 (7.67) |
| BCSS Delayed Recall | 100.2 (15.02) | 100.0 (18.36) | 100.3 (11.69) |
| CES-D (score) | 4.3 (3.47) | 4.7 (3.59) | 3.8 (3.60) |
| EyeAGE L (yrs) | 56.0 (2.54) | 56.4 (1.92) | 55.5 (3.24) |
| EyeAGE R (yrs) | 56.4 (2.67) | 56.8 (1.94) | 55.9 (3.47) |

Safety measures during the screening/baseline visit, every two months during the 6-month intervention period and the 6-month follow-up visit included: 1) Comprehensive Metabolic Panel (CMP), 2) thyroid stimulating hormone (TSH), 3) FSH, 4) MSQ, 5) Resting Systolic (SPB) and 6) Diastolic Blood Pressure (DPB), 7) alanine aminotransferase (ALT), 8) alkaline phosphatase (ALP), 9) Lymphocyte %, 10) Mean Cell Volume (MCV), 11) Red Cell Distribution Width (RDW%), 12) Alkaline Phosphatase (ALK), 13) White Blood Cells (WBC) and blood platelet count. The ECG was conducted at baseline, 4-month and 6-month post intervention clinic visits. In-home procedures included self-reporting any symptoms, issues, or problems with the treatment procedures to the study coordinator by phone. Quarterly safety reports including symptoms, adverse events and withdrawals were submitted to the Data Safety Monitoring Board as per the IRB-approved protocol.

Metabolic function, e.g., fasting glucose and insulin; IR by Homeostatic Model [fasting glucose (mmol/L) x fasting insulin (mIU/L)/22.5 (HOMA-IR)], HbA1C, TC, LDL HDL, triglycerides and phenotypic age [Albumin, creatinine, glucose, C-reactive protein (CRP), Lymphocyte %, MCV, RDW%, ALK, WBC, age (years)] were measured using samples of whole blood obtained during the screening/baseline visit in a certified laboratory and again during the 6-month follow-up visit. For all blood tests, venous blood was collected in the morning after a 12-hour fast with the study participant seated. Tubes containing 1.5 mg/ml EDTA were used for procedures requiring plasma, in tubes with no additives for serum measures and in citrate tubes for homeostasis endpoints.

Fat oxidation by RQ was measured at rest under standardized conditions with the study participant lying supine on a bed following a 12-hour fast and a 30-minute resting period.^31^ Metabolic measurements were continuously taken during the test. Expired gases were sampled and analyzed for VO_2_ and VCO_2_ concentrations by indirect calorimetry calculated via a metabolic cart (Cosmed Q-NRG, Rome, Italy) using a canopy hood set up (Cosmed Q-NRG, Rome Italy). The RQ was evaluated as an index of fat oxidation.

Depression was measured by the Center for Epidemiologic Studies Depression Scale (CES-D Scale) CES-D Scale.^32^ Study participants completed the 20-item scale indicating how they felt or behaved during the past week. Responses were totaled by the test administrator. Possible scores range from 0 to 60, with higher scores indicating the presence of more depressive symptomatology. A score of 16 or higher represents a clinical diagnosis of depression.

Cardiorespiratory Fitness (CRF) was assessed by measuring Submaximal Oxygen Uptake (VO_2_) during a graded treadmill test using a modified Balke protocol to determine CRF.^33^ A warm-up session of approximately 5 minutes established a comfortable walking speed. Study participants then walked on the treadmill for a 2-minute warm-up at 0% grade. The grade of the treadmill was then increased by 2.5% every 2 minutes until the participant could no longer continue or achieved 80% of age-predicted maximal heart rate. During the test, VO_2_ and carbon dioxide (VCO_2_) production, ventilation (VE) and respiratory exchange ratio (RER) were calculated via a metabolic cart (Cosmed Quark CPET, Rome, Italy) and a two-way non-re-breathing mask (Cosmed K2 Mask, Rome, Italy).

Total body weight and height were measured with an electronically calibrated scale and stadiometer (Pelstart Health-o-meter Professional 500KL) with participants in light, loose fitting clothing without shoes at baseline, 2-, 4- and 6 months. Waist circumference was measured in study participants with a tape measure at the umbilicus at the end of a normal expiration by trained research staff at baseline and 6-month follow-up.

Dual energy x-ray absorptiometry (DEXA) was used to determine body composition [%fat, LM, FM, BMD, bone mineral content (BMC)] and central adiposity [(Visceral Adipose Tissue area volume and mass (VAT)]. A Hologic Horizon A DEXA device measured study participants at baseline and 6-month follow-up. Study participants were instructed to remove all metal and jewelry including their shoes before lying down on their back on the scanning table with their arms to their side.

Caloric Intake (CI) was assessed using the National Institutes of Health/National Cancer Institute ASA24-2020 Dietary Assessment Tool^34^ via a self-administered 24hr food recall at baseline and 6-month follow-up.

The eyeAge retinal age gap images were obtained by retina scan (Eidon Af True Color Confocal Scanner) via standard clinical procedures in the left and right eye of the study participant at baseline and 6-month follow-up. Images were transported to the Kapahi lab for analysis using the previously reported general aging clock procedures.^35^

Neuromuscular Function was measured by assessing participants’ PF using the short physical performance battery.^36^ Study participants completed three performance measures including the standing balance test, the 4-meter walking speed test, and the chair stand test at baseline and 6-month follow-up. Each performance measure was scored from 1–4 points and totaled to calculate the final PF score.

Cognitive function was measured per standard protocol with the cut-off of 26 by the Mini-Mental State Examination (MMSE).^37^ The MMSE is comprised of 30 questions that assesses cognitive function by evaluating attention and orientation, memory, registration, recall, calculation, language and ability to draw a complex polygon. The immediate (IM) and delayed recall test was administered to the study participants using the Advanced Digital Cognitive Assessment (BrainCheck)^38^ at baseline and 6-month follow-up. BrainCheck is a tablet-based version of six neurocognitive tests, which can be completed in 30 minutes. For the IM, study participants were asked to identify a distractor or target word after 10 words that are displayed in a series.

Procedures for monitoring subject adherence: During 2-, 4-, and 6-month visits, trained clinical research staff interviewed participants about supplement intake, forgetfulness, and barriers to adherence. Staff reviewed supplement details, addressed side effects and concerns, and answered questions. Monitoring via pill counts were additional indicators of adherence.

### 2.9 Statistical Methods

Sample size estimates were not conducted due to the one-group, non-comparer pilot study design with a major focus of examining safety, while observing changes in primary and secondary outcomes. The primary analysis followed an intent-to-treat (ITT) approach, including all participants. For those who were withdrawn or lost to follow-up, all available data were incorporated into the ITT analysis. A secondary per-protocol analysis was conducted among participants who completed the study. Descriptive statistics were reported as means and standard deviations at baseline and follow-up. Changes in study endpoints over time were assessed using linear mixed-effects models, which account for within-subject correlations and do not require imputation for missing data. Endpoint measurements at baseline and follow-up were used as dependent variables, while time (baseline vs. follow-up) was included as the independent variable to estimate longitudinal changes. The models incorporated random intercepts (for participants) and random slopes (for time) to account for individual variability. An unstructured correlation matrix was used to model time dependencies. The normality of residuals was assessed via QQ plots. All statistical analyses were conducted using SAS version 9.4 (SAS Institute Inc.).

## 3 Results

The study began recruitment on 27 July 2023 and ended recruitment on 20 November 2023. The final study 6-month post-assessment visit was conducted on 31 MAY 2024 (Figure 1). Outcomes from participants enrolled in the study, including both completers (n=7) and non-completers (n=6), were summarized using descriptive statistics (Table 2). Intent-to-treat analysis showed minimal alterations in safety measures following the six-month intervention (Table 3–5; Figure 1). Safety assessments, RDW% (p=0.009), MCV (p=0.0004) and HDL (p= 0.044), were significantly reduced (Table 3–5). Conversely, depressive symptoms (p=0.002; Figure 3c) increased significantly; however, individual 6-month follow-up scores did not reach the established threshold for the diagnosis of depression (Tables 4–5). No severe adverse events were self-reported or shown in the MSQ during the safety follow-up visits. Six participants reported mild to moderate adverse events (acid indigestion), dissatisfaction with study weight loss results, unrelated symptoms and were lost to follow-up (Table 3). Self-report of intake and pill counts indicated that adherence to the prescribed oral supplement regimen was maintained among those attending the safety and follow-up visits (Table 3).

**Table 3:**
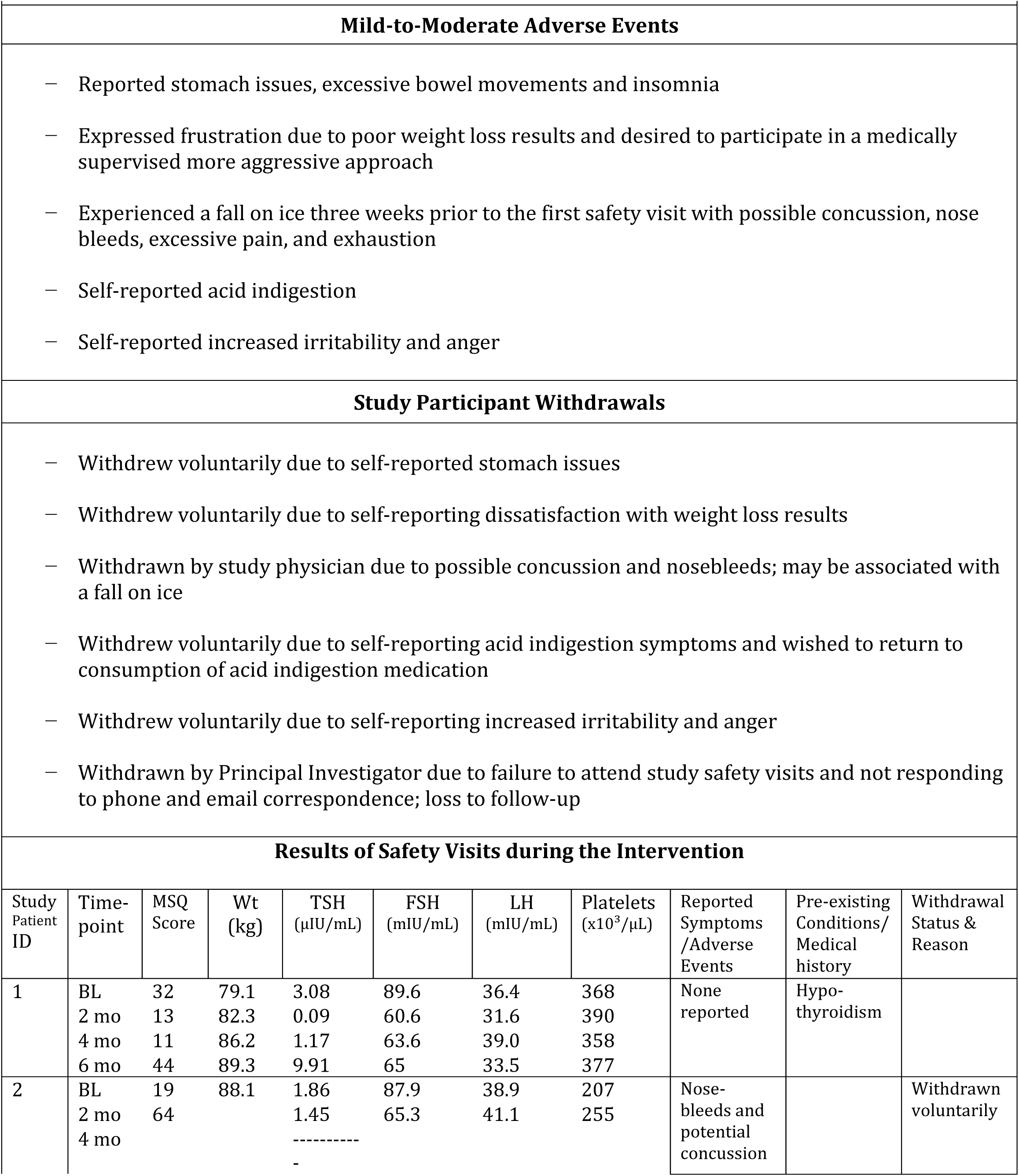
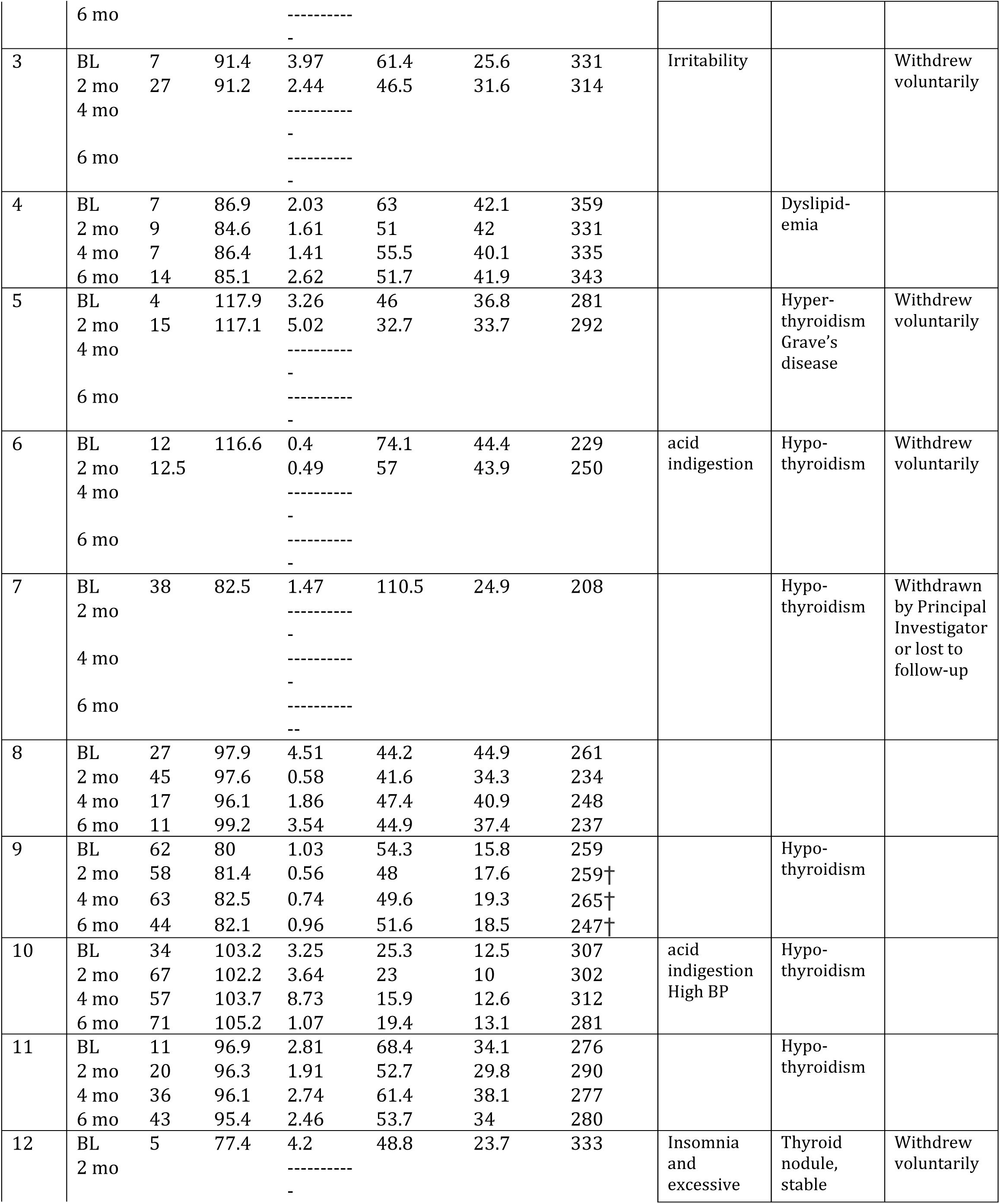
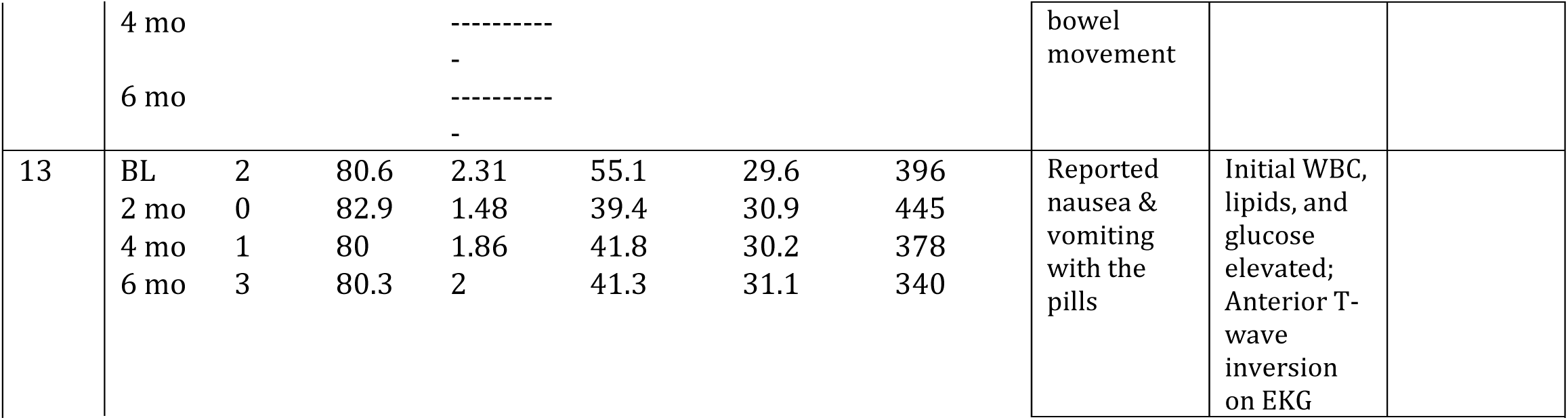
Safety and Tolerability of a Natural, Multi-modal Oral Therapeutic in Study Participants. †Mean platelet volume (MPV) was 8.7 (borderline low, stable) at the 2-month and 4-month safety checks. ID (Participant Identification Number) BL (baseline); MSQ (Medical Symptoms Questionnaire); Wt (Weight); TSH (Thyroid Stimulating Hormone); FSH (Follicle Stimulating Hormone); LH (Luteinizing Hormone); mo (month); BP (Blood Pressure); EKG (Electrocardiogram)

**Table 4:** Change following Daily Oral Supplementation with a Natural, Multi-modal Therapeutic [Intent-to-treat analysis (mixed-effect models)] SD (standard deviation); IQR (Interquartile Range); BMI [Body Mass Index = total body weight (kg)/Height (m)^2^ (BMI)]; SPB (Systolic Blood Pressure); DBP (Diastolic Blood Pressure; BMD (Bone Mineral Density); BMC (Bone Mineral Content); VAT (Visceral Adipose Tissue); HOMA-IR (Homeostatic Model for Insulin Resistance); hbA1C (glycosylated hemoglobin); TG (Triglycerides); TC (total cholesterol); HDL (High Density Lipoprotein); LDL (Low Density Lipoprotein); CRP (C-Reactive Protein); Alb (Albumin); RDW (Red Cell Distribution Width); LYM (Lymphocytes); CREAT (Creatine); MCV (Mean Cell Volume); ALP (Alkaline Phosphatase); WBC (White Blood Cells); RQ (Respiratory Quotient); Submax [Submaximal (80% heart rate max)] VO_2_ (Oxygen Uptake); MMSE (Mini Mental State Examination of Cognitive Function); BCCM (BrainCheck Cognitive Measurement); BCSS (BrainCheck subscale); CES-D (Center for Epidemiologic Studies Depression Scale); eyeAge (Retina Scan: General Aging Clock Retina Imaging)

| Change Following Daily Oral Supplementation with a Natural, Multi-modal Therapeutic |  |  |
| --- | --- | --- |
| Variables | Change (SD) | p-value |
| Weight (kg) | 1.1 (1.35) | 0.4319 |
| Height (cm) | 1.5 (0.32) | 0.0032 |
| BMI | -0.2 (0.52) | 0.7587 |
| SBP | -5.3 (4.89) | 0.3237 |
| DBP | -2.3 (3.75) | 0.5663 |
| Waist Circumference (cm) | -5.1 (1.57) | 0.0167 |
| Physical Function | 0.4 (0.21) | 0.1420 |
| Fat% | -0.6 (0.48) | 0.2353 |
| Fat Mass (kg) | -0.05 (0.96) | 0.9612 |
| Lean Mass (kg) | 1.0 (0.62) | 0.1552 |
| BMD (g/cm<sup>2</sup>) | 0.01 (0.01) | 0.0188 |
| BMC (g) | 15.2 (23.37) | 0.5397 |
| VAT Area (cm<sup>2</sup>) | 0.6 (8.38) | 0.9484 |
| VAT Volume (cm<sup>3</sup>) | 0.8 (43.43) | 0.9857 |
| VAT Mass (g) | 0.5 (40.23) | 0.9899 |
| HOMA-IR | 0.03 (0.21) | 0.8848 |
| hbA1C (%) | 0.1 (0.13) | 0.3799 |
| FSH ( mIU/mL) | -13.1 (2.47) | 0.0018 |
| TG (mg/dL) | 31.8 (14.20) | 0.0663 |
| TC (mg/dL) | -16.1 (16.39) | 0.3631 |
| HDL (mg/dL) | -9.7 (3.82) | 0.0442 |
| LDL (mg/dL) | -17.0 (10.18) | 0.1451 |
| TSH (uIU/mL) | 0.8 (1.12) | 0.5484 |
| CRP (mg/dL) | -0.1 (0.07) | 0.2401 |
| Alb (g/dL) | -0.2 (0.12) | 0.1108 |
| RDW (%) | -0.2 (0.05) | 0.0094 |
| LYM (%) | 0.9 (0.81) | 0.3302 |
| CREAT (mg/dL) | -0.02 (0.06) | 0.7395 |
| MCV (fL) | -2.3 (0.33) | 0.0004 |
| ALP (U/L) | -7.5 (5.82) | 0.2426 |
| WBC (K/uL) | 0.3 (0.32) | 0.4570 |
| Phenotypic Age | -2.1 (0.87) | 0.0559 |
| RQ | -0.1 (0.02) | 0.0178 |
| Submax VO<sub>2</sub> (ml/min) | 78.2 (84.61) | 0.3907 |
| Submax VO<sub>2</sub>/kg (ml/min/kg) | 1.1 (0.89) | 0.2514 |
| Lean Adjusted Submax VO<sub>2</sub> | 0.8 (1.48) | 0.5979 |
| RQ during exercise | 0.1 (0.02) | 0.0173 |
| Caloric Intake | -384.5 (105.15) | 0.0106 |
| MMSE Total | 3.8 (8.49) | 0.7049 |
| BCCM Total | 2.5 (3.75) | 0.5315 |
| BCSS Trails A | 2.0 (3.62) | 0.6096 |
| BCSS Trails B | 1.2 (2.79) | 0.6897 |
| BCSS Digit | -17.2 (11.63) | 0.1900 |
| BCCS Stroop | -2.8 (3.33) | 0.4251 |
| BCCS Immediate Recall | 10.6 (5 4.10) | 0.0422 |
| BCSS Delayed Recall | 9.7 (5.16) | 0.1080 |
| CES-D | 4.0 (0.75) | 0.0019 |
| EyeAGE L (yrs) | 2.0 (1.14) | 0.1257 |
| EyeAGE R (yrs) | 0.9 (0.74) | 0.2899 |

**Table 5:** Change following Daily Oral Supplementation with a Natural, Multi-modal Therapeutic (mixed-effect models without intent-to-treat analysis) SD (standard deviation); IQR (Interquartile Range); BMI [Body Mass Index = total body weight (kg)/Height (m)^2^ (BMI)]; SPB (Systolic Blood Pressure); DBP (Diastolic Blood Pressure; BMD (Bone Mineral Density); BMC (Bone Mineral Content); VAT (Visceral Adipose Tissue); HOMA-IR (Homeostatic Model for Insulin Resistance); hbA1C (glycosylated hemoglobin); TG (Triglycerides); TC (total cholesterol); HDL (High Density Lipoprotein); LDL (Low Density Lipoprotein); CRP (C-Reactive Protein); Alb (Albumin); RDW (Red Cell Distribution Width); LYM (Lymphocytes); CREAT (Creatine); MCV (Mean Cell Volume); ALP (Alkaline Phosphatase); WBC (White Blood Cells); RQ (Respiratory Quotient); Submax [Submaximal (80% heart rate maximum)] VO_2_ (Oxygen Uptake); MMSE (Mini Mental State Examination of Cognitive Function); BCCM (BrainCheck Cognitive Measurement); BCSS (BrainCheck subscale); CES-D (Center for Epidemiologic Studies Depression Scale); eyeAge (Retina Scan:General Aging Clock Retina Imaging)

| Change following Daily Oral Supplementation with a Natural, Multi-modal Therapeutic |  |  |
| --- | --- | --- |
| Variables | Change (SD) | p-value |
| Weight (kg) | 1.6 (1.34) | 0.2783 |
| Height (cm) | 1.6 (0.34) | 0.0037 |
| BMI | -0.02 (0.52) | 0.9642 |
| SBP | -3.6 (4.98) | 0.4921 |
| DBP | -0.9 (3.87) | 0.8322 |
| Waist Circumference (cm) | -4.8 (1.43) | 0.0153 |
| Physical Function | 0.1(0.24) | 0.5757 |
| FAT% | 0.2 (0.45) | 0.7398 |
| Fat Mass (kg) | 0.8 (0.88) | 0.3892 |
| Lean Mass (kg) | 1.0 (0.62) | 0.1583 |
| BMD (g/cm<sup>2</sup>) | 0.02 (0.01) | 0.0197 |
| BMC (g) | 12.8 (23.50) | 0.6051 |
| Vat Area (cm<sup>2</sup>) | 1.7 (8.31) | 0.8434 |
| VAT Volume (cm<sup>3</sup>) | 6.7 (43.09) | 0.8813 |
| Vat Mass (g) | 6.0 (39.92) | 0.8855 |
| Fasting Insulin (mU/L) | 1.4 (0.72) | 0.0997 |
| HOMA-IR | 0.01 (0.20) | 0.9605 |
| hbA1C (%) | -0.01 (0.10) | 0.9432 |
| FSH ( mlU/mL) | -11.1 (2.72) | 0.0094 |
| Fasting TG (mg/dL) | 23.6 (16.05) | 0.2008 |
| TC (mg/dL) | -18.6 (17.08) | 0.3268 |
| HDL (mg/dL) | -5.6 (3.88) | 0.2067 |
| LDL (mg/dL) | -12.7 (10.55) | 0.2817 |
| TSH (ulU/mL) | 0.5 (1.09) | 0.7032 |
| CRP (mg/dL) | -0.04 (0.09) | 0.6797 |
| Alb (g/dL) | -0.4 (0.17) | 0.0662 |
| RDW (%) | -0.2 (0.05) | 0.0181 |
| LYM (%) | 0.9 (0.81) | 0.3354 |
| CREAT (mg/dL) | -0.1 (0.07) | 0.1879 |
| MCV ( fL) | -2.3 (0.33) | 0.0008 |
| ALP (U/L) | -8.2 (5.63) | 0.2038 |
| WBC (K/uL) | 0.5 (0.35) | 0.1835 |
| Phenotypic Age | -2.7 (0.97) | 0.0373 |
| RQ | -0.1 (0.02) | 0.0169 |
| Submax VO<sub>2</sub> (ml/min) | 99.6 (101.22) | 0.3633 |
| Submax VO<sub>2</sub>/kg (ml/min/kg) | 1.1 (1.08) | 0.3636 |
| Lean Adj Submax VO<sub>2</sub> | 1.5 (1.93) | 0.4641 |
| RQ during exercise | 3.0 (1.99) | 0.1824 |
| Caloric Intake | -356.0 (111.14) | 0.0185 |
| MMSE Total | 3.4 (8.49) | 0.7049 |
| BCCM Total | 2.4 (4.12) | 0.5775 |
| BCSS Trails A | 1.7 (3.62) | 0.6529 |
| BCSS Trails B | 0.3 (2.91) | 0.9189 |
| BCSS Digit | -16.9 (11.99) | 0.2094 |
| BCSS Stroop | -1.9 (3.51) | 0.6161 |
| BCSS Immediate Recall | 10.0 (4.66) | 0.0755 |
| BCSS Delayed Immediate Recall | 9.9 (6.35) | 0.1713 |
| CES-D | 4.0 (0.76) | 0.0018 |
| EyeAGE L (yrs) | 1.5 (1.13) | 0.2427 |
| EyeAGE R (yrs) | 0.6 (0.73) | 0.4470 |

Significant reductions were observed in the primary outcomes, RQ (p=0.02; Figure 2a) and waist circumference (p=0.02; Figure 3a), as well as the secondary outcomes FSH (p=0.002; Figure 2d) and CI [(p=0.01; Figure 3b) among study participants (n=7; intent-to-treat mixed-effects models; Tables 4–5). Additionally, significant increases were noted in height (p=0.003; Tables 4–5), BMD (p=0.02; Figure 2b) and IM (p=0.04; Figure 3d) in the participants (n=7; intent-to-treat mixed-effect models; Tables 4–5). Reductions in phenotypic age did not achieve statistical significance in the intent-to-treat analysis (p=0.055; Table 4). However, a significant decline was observed when mixed-effect models were applied without intent-to-treat analysis [(p=0.037; Figure 2c) Table 5]. Changes in BMI (p=0.76), BF% (p=0.23), TC (p=0.36), LDL (p=0.15), CRP (p=0.24) and increases in LM (p=0.155), BMC (p=0.54), PF (p=0.14) and VO2 (p=0.39), did not reach statistical significance (Tables 4–5). No significant changes were observed in HOMA-IR, HbA1c and VAT (Tables 4–5).

**Figure 2.**
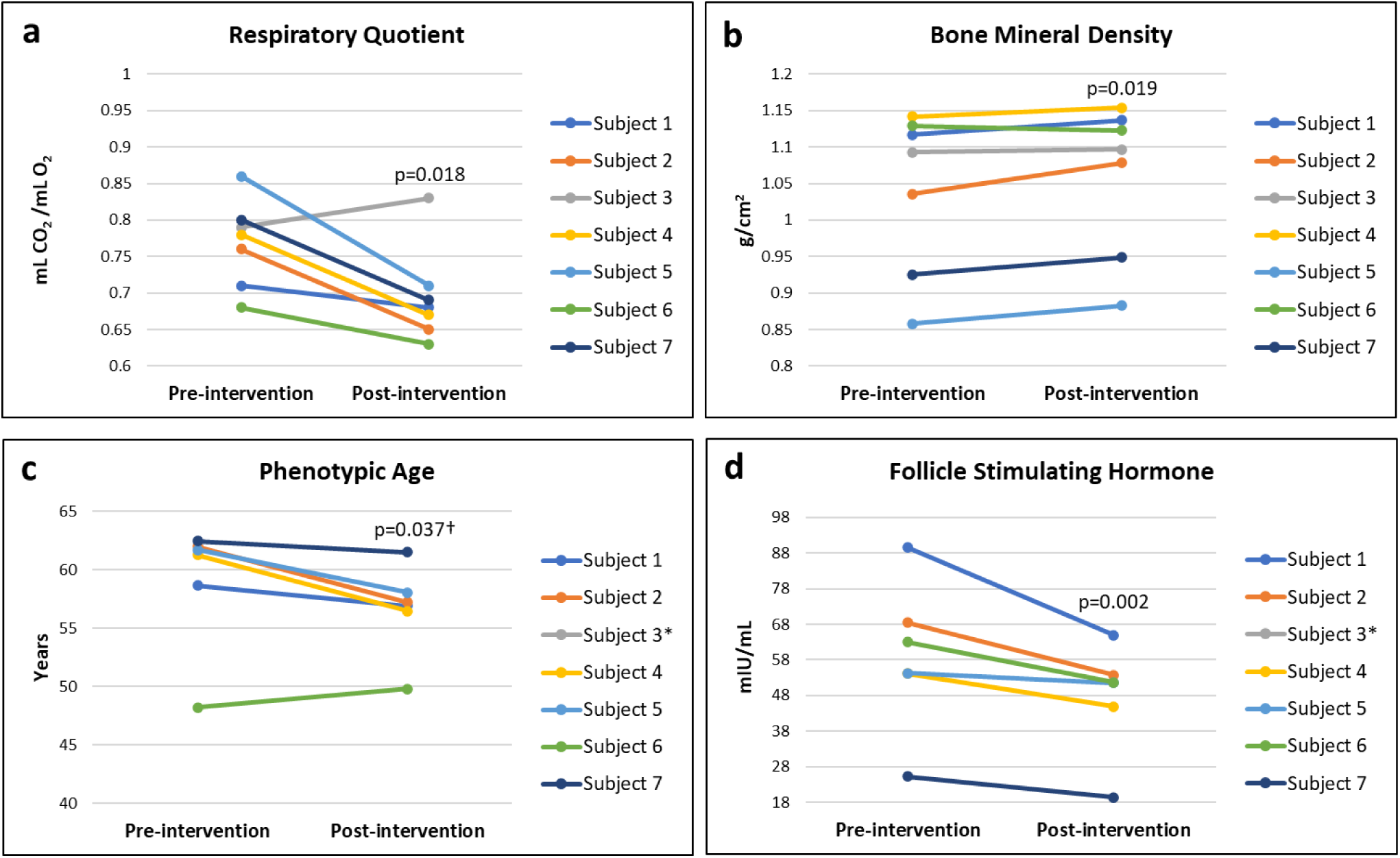
**a**: Change in Respiratory Quotient (RQ) following six months of daily oral supplementation with a natural, multi-modal therapeutic **b**: Change in Bone Mineral Density (BMD) following six months of daily oral supplementation with a natural, multi-modal therapeutic **c**: Change in Phenotypic Age following six months of daily oral supplementation with a natural, multi-modal therapeutic **d**: Change in Follicle Stimulating Hormone (FSH) following six months of daily oral supplementation with a natural, multi-modal therapeutic * Missing lab results † Mixed-effect models

**Figure 3.**
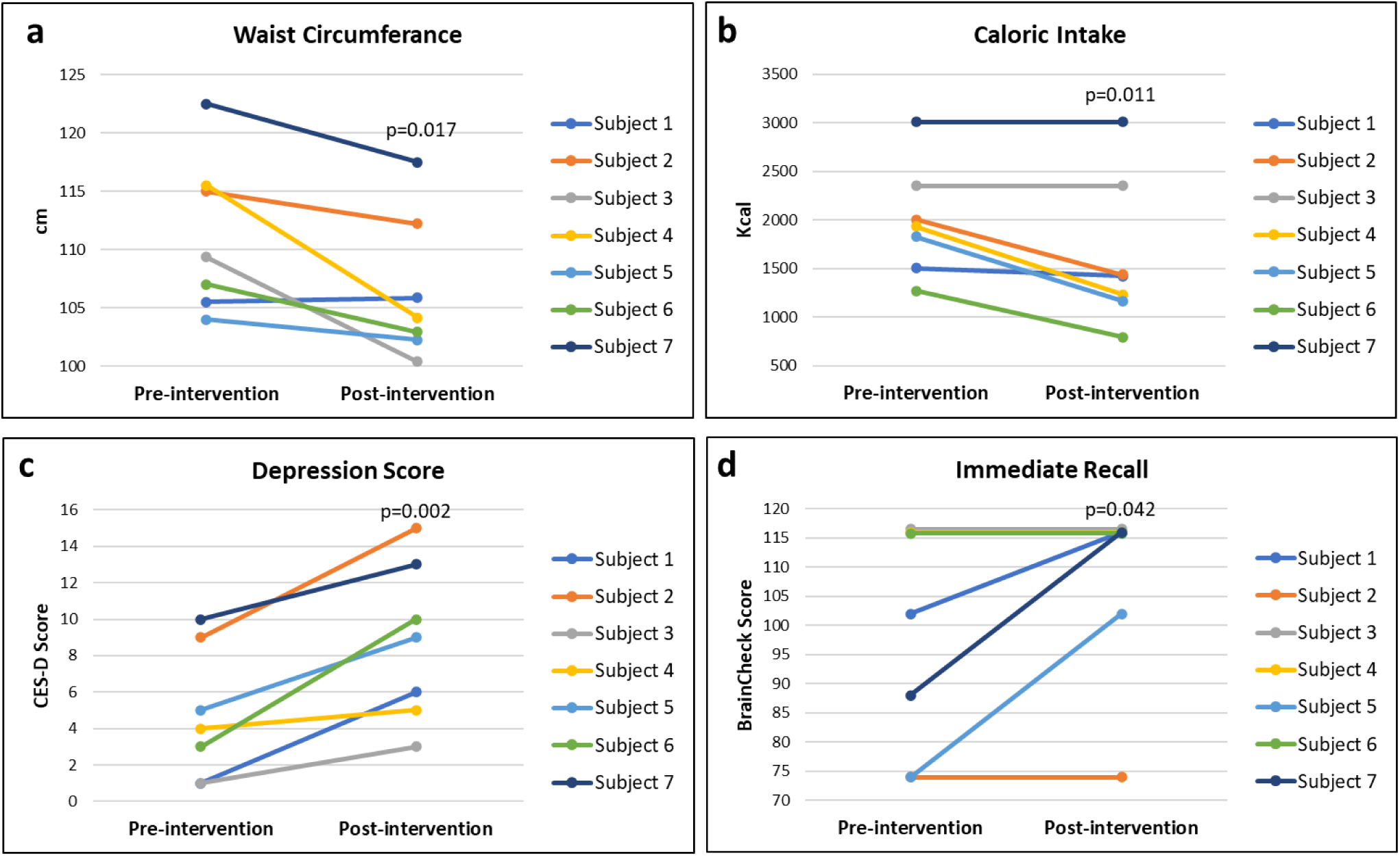
**a:** Change in Waist Circumference following six months of daily oral supplementation with a natural, multi-modal therapeutic **b**: Change Caloric Intake (CI) following six months of daily oral supplementation with a natural, multi-modal therapeutic **c**: Change in Depression (CES-D Score) following six months of daily oral supplementation with a natural, multi-modal therapeutic **d**: Change in Immediate Recall (IM) following six months of daily oral supplementation with a natural, multi-modal therapeutic

## 4 Discussion

Postmenopausal women are at increased risk for complex chronic conditions caused by a cascading array of insults to multiple biological pathways leading to functional declines as they age.^39^ However, the deleterious metabolic, physical and cognitive determinants driving these age-related declines are preventable, manageable and, in some cases, reversible through targeted therapies informed by clinical and translational research.^29,40^ This study represents a pivotal milestone, translating extensive innovative in vitro and in vivo research into a human population. Specifically, a natural, multimodal oral therapeutic developed with safe, well-characterized ingredients, was tested in postmenopausal women based on findings derived from studies in worms, mammalian cell lines, and mice. The results of this trial mark the first report of oral supplementation in humans, paving the way for targeted interventions to improve health outcomes in this vulnerable population.

In this pioneering clinical study, we investigated the safety and observed the effects of a combination of vitamins and natural compounds in postmenopausal women with obesity. The orally-delivered therapeutic is designed to activate multiple longevity-enhancing pathways that address the impacts of vitamin and mineral deficiencies. These deficiencies reduce cell differentiation and proliferation, immunosenescence, and inflammaging,^6,20,41^ which contribute to a shortened healthspan. The therapeutic potential is supported by extensive in vivo and in vitro studies, which demonstrate its ability to improve metabolic and brain health, reduce FSH levels, and enhance both healthspan and lifespan.^20,22–24^ Prior preclinical findings helped identify a unique phenotype of aging-related debility, particularly in older women. Using an integrated, precision medicine approach, it was further confirmed that postmenopausal women would be most likely to benefit from the oral supplementation. As such, findings from this pilot study represent a translational application of recent geroscience recommendations in a clinical health care setting.^42^

In this first-in-human pilot study, multiple pathways that enhance longevity were engaged through the use of several vitamins and natural compounds. These compounds functioned synergistically to significantly increase fat oxidation, height, BMD, and IM in parallel with reductions in waist circumference, FSH, CI and phenotypic age. These results not only corroborate previous pre-clinical findings but also provide novel insights into the role of glycation reduction in fat oxidation, FSH, and phenotypic age. Conversely, HOMA-IR and HBA1c were not significantly altered. However, overall results remain relevant as older women especially demonstrate a limited ability to adopt and adhere to the healthy diet and PA behaviors^43^ integral to maintaining metabolic, physical and cognitive function. The complexity of obesity-related aging frailty further compounds healthy behavior adherence due to multifactorial molecular, biological and socio-environmental determinants.^28^ As such, natural oral supplementation offers an attainable, convenient and easily accessible therapeutic for postmenopausal women with obesity at increased risk of aging-related nutrient deficiencies.

The natural, multimodal therapeutic was shown to be safe and generally well-tolerated in the majority of study participants in this outpatient, clinical setting. Adverse events were mild to moderate but, in some participants, resulted in self-withdrawal from the study. The medical history of six of the participants indicated the presence of hypothyroidism and one with hyperthyroidism (Table 2). Three of these participants withdrew from the study including the one with hyperthyroidism. However, four completed the study and two of these continued despite reporting acid indigestion symptoms. Additional research is needed to examine the mediating potential of thyroid disease to study outcomes, especially in postmenopausal women due to an increased risk of impaired bone metabolism.^44^ Moreover, increases in depression scores, which were statistically but not clinically significant, were observed, which may indicate that the participants’ experience was not positive. This could be due to self-reported side effects or the lack of a significant reduction in body weight observed during study visits. Findings indicating that two study participants withdrew to seek alternative weight loss therapies support this concept. Notably, participants were advised to maintain dietary and exercise patterns. Thus, daily oral supplementation may be more efficacious if paired with nutrition and exercise counseling, treatments that would be complimentary to enhancing the overall effect of the oral, multimodal therapeutic.

Fat oxidation objectively measured during rest by indirect calorimetry provides an accurate estimate of fat and carbohydrate substrate utilization (RQ).^45^ The ability to utilize fat (fat oxidation) preferably over carbohydrates at rest is a well-established marker for healthy metabolic function.^45^ Fat oxidation is linearly associated with increased physical activity and fitness^46^ and inversely associated with excess adiposity, VAT and IR.^47^ Numerous publications report significant increases in fat oxidation in adults following exercise training,^48^ but few report improvements following nutrition therapeutics.^49^ Currently, there are no studies examining fat oxidation following nutrition therapeutics in postmenopausal women specifically. And, surprisingly, Guzel and others^50^ reported that 10 weeks of walking exercise was insufficient to improve fat oxidation in postmenopausal women. However, this may be explained by the short duration of training and small sample size. In the current study, fat oxidation was significantly increased after six months of daily, oral supplementation despite the small number of participants completing the study. These are significant findings as recent research proposes that impaired fat oxidation may promote bone marrow adiposity and reductions in BMD and musculoskeletal health in postmenopausal women.^51^ An increase in height was also observed following the six-month intervention. This may be attributable to the increase in BMD and an overall improvement in musculoskeletal health, supporting the functionality of postural muscles and aiding in a more upright posture.^52^ Additionally, the improvement in BMD may have helped reduce kyphosis, a condition commonly associated with height loss in postmenopausal women.^52^ These factors suggest that increasing BMD can improve both bone health and posture in postmenopausal women. Bone mineral density in postmenopausal women is well-studied with numerous clinical trials reporting improvements following pharmacologic therapies^53^ albeit with numerous side effects. Less studied and reported are direct associations between vitamin deficiencies and reduced bone mineral density,^54^ and improved BMD following supplementation such as with resveratrol.^55^ Interestingly, prior studies show that regular supplementation with low-dose resveratrol also enhances cognition and cerebrovascular function in postmenopausal women.^11^ Likewise, significant increases in IM, a subscale of cognitive function, were also observed in the current study coinciding with significant increases in BMD. Additional studies are needed to confirm positive BMD and cognitive outcomes following treatment with the natural, multimodal therapeutic in postmenopausal women who are at increased risk for bone loss, especially in those with chronic conditions such as metabolic, cardiovascular and thyroid disease.^44^

Studies examining phenotypic age alterations in postmenopausal women specifically are limited. In the current pilot investigation, we observed a significant reduction in phenotypic age. However, once intent-to-treat analysis was applied, results did not achieve significance. Entering menopause early or late was recently shown to be associated with a higher phenotypic age, findings that were attenuated with positive lifestyle behaviors including maintenance of a healthy body weight.^56^ Additional reports confirmed these epidemiological findings in adults, 20 years and older.^57^ Clinical trials examining the effect of nutrition supplementation on phenotypic age in postmenopausal women with obesity are essential to understanding aging-related mechanisms.

Analogous to the preclinical findings in mice, the current study results showed a significant reduction in FSH following six months of treatment in postmenopausal women with obesity. Central adiposity measured by waist circumference was similarly reduced. Previously, obesity was shown to be linearly associated with FSH in postmenopausal, but not premenopausal women.^54^ In a summary of epidemiological studies, Mao and others concluded that FSH may promote abnormal fat metabolism and cognitive impairment in menopausal women. Conversely, a large scale epidemiological analysis of US NHANES data showed that increased FSH levels were linked to decreased risk for metabolic syndrome in postmenopausal women.^58^ Comparably, Wang and others reported that obesity prevalence, diabetes mellitus, and metabolic syndrome also decreased with increasing quartiles of FSH in 2658 postmenopausal women.^59^ Klisic and colleagues reported that elevated waist circumference predicted lower FSH in a cohort of 50 postmenopausal women with healthy weight status, and 100 with overweight or obesity.^60^ Interestingly, although findings from the current study show significant reductions in both fat oxidation and waist circumference, Karppinen and others recently reported that FSH was not significantly associated with changes in fat oxidation in a combined cohort of 42 pre-to-postmenopausal women, 52-58 years of age.^46^ Mixed findings concerning the benefits of FSH in postmenopausal in both clinical trials and epidemiological studies, and in the current study warrant further investigation.

Recently technologies using retina imaging illuminate strong relationships between IR,^61^ CO and mortality.^62^ Buck Institute investigators showed that eyeAge predicted chronological age with a mean absolute error of 3 yrs. Results maintained relatively stable accuracy in longitudinal testing of these individuals. Thus, an increased retinal age gap,^62^ which represented the difference between the eyeAge and the known chronological age equated to an earlier mortality. In the current study, eyeAge was not significantly altered following the treatment. However, this may be attributable to the short treatment period and small sample size. Larger clinical trials are needed to examine the effect of this natural, multimodal therapeutic on eyeAge.

### 4.1 Limitations

The current findings are limited by the small sample size and lack of a placebo control. Non-significant results may be due to insufficient statistical power; 6 participants (46%) did not complete the study. Difficulty meeting enrollment targets may be attributed to the strict exclusion criteria. In addition, although the intervention promoted no severe adverse events, 46% were loss to follow-up primarily due to reports that the product was not well tolerated. However, participants lost to follow-up were compared to those with complete data. An intent-to-treat analysis was employed and results were similar with the exception of phenotypic age findings. Regardless, several significant beneficial health impacts were observed in postmenopausal women with obesity following six months of daily, oral supplementation with the natural, multimodal therapeutic. More importantly, no severe adverse events were reported during the clinical trial.

### 4.2 Conclusion

This study represents the first clinical trial to translate preclinical findings from a natural, oral multimodal therapeutic into a human population. Preliminary results indicate that the intervention is safe and generally well-tolerated in postmenopausal women with obesity. Significant improvements were observed in fat oxidation, BMD and IM, a subscale of cognitive function, while waist circumference, FSH, CI and phenotypic age were reduced following the intervention. Sub-max VO_2_ and PF were not altered, which may indicate that participants did not increase exercise activity. Thus, the therapeutic may operate autonomously to improve aging-related metabolic and biological outcomes. Moreover, no significant alterations in IR or HbA1C were observed indicating that six months of oral supplementation may be insufficient to positively alter glucose control or maintenance. Hence, the decrease in resting RQ may be the result of a shift in metabolic substrate utilization independent of insulin homeostasis. Notably, average baseline values of HbA1C were within normal limits. Also, volunteers with T2DM or prescribed medications for IR were excluded. As such, it is unknown if a similar treatment intervention will promote improvements in HbA1C in those individuals. Significant decreases in HDL, RDW% and MCV and increased depression scores also warrant further investigation in future randomized-controlled clinical trials. Nonetheless, the findings from this pilot study provide preliminary evidence that a natural, multimodal therapeutic is safe in humans and demonstrate preliminary effectiveness toward positively altering several obesity-related aging pathologies. The utilization of retinal imaging technology (eyeAge) represents an innovative advancement in geroscience research.^35^ Moreover, translation of our findings into clinical practice can be immediate as the oral therapeutic is widely available and affordable, which is contrary to currently-available pharmacologic-based weight loss medications.^10^ Future research should include double-blind, randomized-controlled trials to confirm these results in a larger population of postmenopausal women with obesity. Younger women with overweight conditions should be included to explore the potential of the natural, multimodal therapeutic toward preventing aging-related metabolic, cognitive and functional decline.

## Data Availability

All data produced in the present study are available upon reasonable request to the authors.

## Acknowledgments

The author(s) thank the medical and laboratory staff at the Hoskinson Health and Wellness Clinic, Gillette, WY, USA for providing assistance with study assessment and intervention procedures, which were funded by the Hoskinson Health and Wellness Clinic, Gillette, WY, USA.

## Author Contributions

TT: Conceptualization, Methodology, Data Curation, Investigation, Project Administration, Visualization, Writing – review & editing; VT: Conceptualization, Methodology, Writing – review & editing; WH: Resources, Supervision, Writing - review & editing; KP: Data Curation, Investigation, Writing – review & editing; MS: Conceptualization, Methodology, Visualization, Writing – original draft; CLM: Conceptualization, Writing – review & editing; CM: Formal Analysis, Software, Writing – review & editing; PK: Conceptualization, Writing – review & editing; SD: Project Administration, Supervision, Writing – review & editing

## Conflict of Interest

TT, KP, MS and SD are employed by the study sponsor, the Hoskinson Health and Wellness Clinic. Author CM is a paid consultant for the Hoskinson Health and Wellness Clinic. Author WH has a financial interest in the Hoskinson Health and Wellness Clinic. Author PK has a financial interest in Juvify, LLC.

## Funding

Research relating to this manuscript was supported by the study sponsor, Hoskinson Health and Wellness Clinic, and Juvify, LLC.

## Data Availability Statement

Data available on request from the authors:

The data that support the findings of this study are available from the corresponding author upon reasonable request.

## Ethics Approval Statement

Study procedures were completed with approval and oversight by the Biomedical Research Alliance of New York (BRANY) Institutional Review Board (IRB). This study conforms to recognized standards by US Federal Policy for the Protection of Human Subjects.

## Patient Consent Statement

N/A

## Permission To Reproduce Material From Other Sources

N/A

## Clinical Trial Registration

Clinicaltrials.gov #NCT06242535

The final completion of the clinicaltrials.gov registration for this study was slightly delayed due to technical issues with the online registration. This resulted in delays receiving the PRS number for our newly registered organization, the Hoskinson Health and Wellness Clinic, located in a rural community in Wyoming, United States. Thus, we enrolled participants a few weeks prior to the clinicaltrials.gov registration date posted on the site.

## Notes

### Clinical Trial

NCT06242535

### Author Declarations

IRB of Biomedical Research Alliance of New York gave ethical approval for this work.

